# Maternal Functional Difficulty and Child Undernutrition in Malawi: A Nationally Representative Cross-Sectional Study(2024 DHS)

**DOI:** 10.64898/2026.08.03.26359624

**Authors:** George Chikondi Samu, Rubina Karki, Noreen Clara Ng’ambi, Modupe Mary Abiona, Sharada Karki, Sellina Samu, Bishnu Kumar Karki, Gibson Mphemvu

**Affiliations:** School of Public Health, Department of Nutrition and Food Hygiene, Southern Medical University, Guangzhou, China; SureCare Medical Supplies, Lilongwe, Malawi; Department of Nutrition and Dietetics, CAFODAT College, Faculty of Science and Technology, Purbanchal University, Kathmandu, Nepal; MASM Medi Clinics, Lilongwe, Malawi; Ministry of Home Affairs, Government of Nepal, Kathmandu, Nepal; Ambassadors Charity Trust

**Keywords:** Maternal disability, functional difficulty, child undernutrition, stunting, wasting, underweight, Malawi, Demographic and Health Survey, Washington Group

## Abstract

**Background:** Maternal disability can reduce the ability to provide adequate care and increase the likelihood of undernutrition among children. In Malawi, over one third of under-five children are affected by stunting. There has been no study that assessed the association between maternal functional difficulty and undernutrition among children in Malawi. The objective was to assess if maternal functional difficulty is associated with stunting, underweight, and wasting in children in Malawi.

**Methods:** Data were extracted from the nationally representative cross-sectional Malawi 2024 Demographic and Health Survey (MDHS). Mother-child dyads were included for mothers aged 15-49 years and children aged ≤ 59 months, with valid anthropometric z-scores. Maternal functional difficulty was assessed using the Washington Group Short Set on six domains (seeing, hearing, walking, cognition, self-care, communication) and classified as severe vs. no difficulty. Child undernutrition was defined as height-for-age z-score (HAZ) < −2 (stunting), weight-for-age z-score (WAZ) < −2 (underweight), and weight-for-height z-score (WHZ) < −2 (wasting). Survey-adjusted multivariable logistic regression models were fitted, accounting for complex sampling design whilst population attributable fraction was calculated.

**Results:** A total of 4,361 mother-child dyads were analyzed for this study. The overall weighted prevalence of maternal functional difficulty was 3.74% (SE 0.35). There was a reported 36.4% prevalence for child stunting, 8.7% for underweight and 1.7% for wasting. Maternal functional difficulty was not associated with child stunting (AOR 0.85, 95% CI: 0.55-1.33), underweight (AOR 1.06, 95% CI: 0.55-2.02) and wasting (AOR 0.96, 95% CI: 0.26-3.53) after adjustments for confounders. The population-attributable fractions were also found to be negligible for all three nutrition outcomes. Maternal height ≥ 150 cm showed strong protective effect against child stunting (AOR 0.29, 95% CI: 0.16-0.54) as well as belonging to the richest wealth quintile (AOR 0.61, 95% CI: 0.42-0.89).

In conclusion, there was no statistically significant association between maternal functional difficulty and child undernutrition in Malawi. The family and community support networks in Malawi may be buffering the nutritional effects of maternal disability on children. To reduce child undernutrition in Malawi, efforts should focus on maternal height, household wealth and the first 1,000 days interventions.

## Introduction

In low- and middle-income countries, undernutrition ranks top among the common causes of death in children under-five. As at 2022, 148 million, 45 million and 37 million children were stunted, wasted and overweight respectively across the globe^1^. Sub-Saharan Africa and South Asia are the regions with the highest burden of this condition. Here, poverty, food insecurity, recurrent infections and weak health systems sustains this toll across the generations^2^.

Malawi is a low-income country in the southeastern region of Africa and still has high levels of child undernutrition. The 2024 Malawi Demographic and Health Survey (MDHS) reported stunting, underweight, and wasting among children under five years of age at 37%, 9%, and 2%, respectively^3^. These estimates are modest improvements from a poor childhood nutrition situation more than a decade ago, but Malawi is unlikely to achieve the World Health Assembly target of reducing childhood stunting to 15% by 2030^4^. As long as undernutrition remains high in Malawi, thus, there is a need for research on all modifiable risk factors, including those related to maternal health and caregiving ability.

Functional disability refers to the limitation of basic activity for an individual to perform given one or more health conditions^5^. The Washington Group on Disability Statistics has developed a standardized short set of questions that consists of functional difficulty across the six domains, which are seeing, hearing, walking or climbing steps, cognition, self-care, and communication^6^. These were included in the DHS and other nationally representative surveys to allow population studies on disability and its correlates.

Nutrition care may be difficult to provide for children of mothers with functional difficulties. For instance, impaired mobility may prevent her from fetching water, preparing food and access to health facilities^7^. Visual and hearing impairment may inhibit mothers from noticing illnesses or receiving and following guidelines for nutrition. Cognitive impairment may prevent meal planning or responsive feeding. Increased risk of disability is linked with lower education, limited employment and poverty which restricts the unit’s ability to achieve adequate nutrition^8^. Physical difficulties may prove most challenging for women who wish to breast feed^9^.

The available research examining maternal disability and child health, although limited, is slowly expanding. One study conducted using the 2019 Bangladesh Multiple Indicator Cluster Survey data established the relationship between maternal functional difficulty and moderate and severe malnutrition among male children even when controlling for factors such as wealth, education level of the mother and others^10^. A second study using the same dataset was able to establish that children whose mother has functional difficulties, had almost three times higher odds of having functional difficulty as well^11^. However, these studies are based on South Asia, and I was unable to find published literature where the objectives of this study have been carried out in sub-Saharan Africa. The cultural, familial and social systems in Malawi is distinct from South Asia. Hence, it is necessary to explore if the same patterns are evident in Malawian settings.

The aim of this analysis was to investigate the association between maternal functional impairment and child undernutrition (stunting, underweight and wasting) among mother-child dyads in Malawi using the 2024 MDHS data, a nationally representative dataset. Our hypothesis is that the odds of child undernutrition is higher among children with mothers having functional difficulty as compared to those without functional difficulty.

## Methods

### Study Design and Data Source

This was a cross-sectional analysis of the data from the 2024 Malawi Demographic and Health Survey (MDHS), which was conducted by Malawi National Statistical Office (NSO) with technical support from ICF International under the auspices of The DHS Program ^3^. Two-stage stratified cluster sampling was adopted for the survey. In the first stage, out of 2024 Malawi Population and Housing Census sampling frame, 793 enumeration areas (EAs) were selected with probability proportional to size. In the second stage, about 30 households are selected from each selected EA, for a total of 23,095 occupied households. All women aged 15-49 years who were usual members of selected households or stayed the night before the survey are eligible for an interview. Anthropometric measurements were obtained for children aged 0-59 months. The 2024 MDHS used Washington Group Short Set of questions on disability as the core module, which administered to all eligible women. The dataset was obtained with registration process from The DHS Program.

### Study Population

We included mother-child dyads meeting the following criteria: the mother was aged 15-49 years with complete Washington Group Short Set data; the child was under 59 months; the child had valid height-for-age, weight-for-age, and weight-for-height z-scores; and the mother and child could be linked through household and line number variables. Children with flagged z-scores (below −600 or above 600 on the DHS scale) were excluded^12^.

### Variables

Maternal functional difficulty was classified using the Washington Group Short Set six-domain responses. Following standard analytical guidelines, we created a binary indicator of severe functional difficulty (our primary exposure), coded as 1 if the mother reported “a lot of difficulty” or “cannot do at all” in any of the six domains (seeing, hearing, walking, cognition, self-care, communication), and 0 if she reported “no difficulty” in all domains. Mothers reporting “some difficulty” only were excluded from the primary analysis to maintain a clear contrast [6]. For sensitivity analysis, we created an alternative “any difficulty” indicator (some, a lot, or cannot do).

#### Outcomes

Child undernutrition was defined using WHO Child Growth Standards [13]. Stunting: height-for-age z-score (HAZ) less than −2 SD. Underweight: weight-for-age z-score (WAZ) less than −2 SD. Wasting: weight-for-height z-score (WHZ) less than −2 SD.

#### Covariates

Based on previous studies^2,10,13^, we included the following covariates: maternal age (15-19, 20-29, 30-39, 40-49 years); maternal education (no education, primary, secondary, higher/other), maternal height (<145cm, 145-149cm, >=150cm), wealth quintile (poorest, poorer, middle, richer, richest), residence (urban, rural); child age (0-11, 12-23, 24-35, 36-47, 48-59 months), child sex (male, female), birth order (1, 2-3, 4-5, 6+). Marital status was not included in the analysis since all mothers in the analytic sample were married (i.e., a single level).

### Statistical Analysis

We used R version 4.6.0 with the survey package to account for the complex sampling design^14^. We specified cluster (EA), stratification, and sampling weights (household weight divided by 1,000,000). We first described the study population and estimated the weighted prevalence of maternal functional difficulty by domain and of child undernutrition overall and by maternal functional difficulty status. We then fitted three separate survey-adjusted multivariable logistic regression models, one for each undernutrition outcome, reporting adjusted odds ratios (AOR) with 95% confidence intervals (CI). We calculated population attributable fractions using the formula PAF = Pe x (AOR - 1) / AOR^15^. We conducted sensitivity analyses using the alternative “any difficulty” definition, restricting to firstborn children, and restricting to children aged 12-59 months. Statistical significance was set at p < 0.05.

### Ethics

The 2024 MDHS received ethical approval from the Malawi National Health Sciences Research Committee and the ICF Institutional Review Board. Informed consent was obtained from all participants. The dataset was de-identified before release. We registered our study concept with The DHS Program and received approval to access the data.

## Results

### Study Population

The 2024 MDHS PR file contained a total of 98,549 records from 23,095 households. Among the records, we had 22,833 women aged 15-49 with complete disability data and 6,069 children Under 59 months with valid z-scores. After linking the children to the mothers, a total of 4,361 mother-child dyads were available for analysis. Table 1 presents the characteristics of the study population.

**Table 1.** Characteristics of mother-child dyads in the analytic sample (n = 4,361)

| Characteristic | n | % (unweighted) |
| --- | --- | --- |
| **Maternal functional difficulty** |  |  |
| No difficulty | 4,187 | 96.0 |
| Severe difficulty | 174 | 4.0 |
| **Maternal age group** |  |  |
| 15-19 years | 374 | 8.6 |
| 20-29 years | 2,034 | 46.6 |
| 30-39 years | 1,527 | 35.0 |
| 40-49 years | 426 | 9.8 |
| **Maternal education** |  |  |
| No education | 342 | 7.8 |
| Primary | 2,868 | 65.8 |
| Secondary | 1,005 | 23.0 |
| Higher/Other | 146 | 3.3 |
| **Maternal height** |  |  |
| <145 cm | 233 | 5.3 |
| 145-149 cm | 477 | 10.9 |
| >=150 cm | 3,651 | 83.7 |
| **Wealth quintile** |  |  |
| Poorest | 1,079 | 24.7 |
| Poorer | 936 | 21.5 |
| Middle | 871 | 20.0 |
| Richer | 775 | 17.8 |
| Richest | 700 | 16.1 |
| **Residence** |  |  |
| Rural | 3,618 | 83.0 |
| Urban | 743 | 17.0 |
| **Child age group** |  |  |
| 0-11 months | 661 | 15.2 |
| 12-23 months | 917 | 21.0 |
| 24-35 months | 923 | 21.2 |
| 36-47 months | 957 | 21.9 |
| 48-59 months | 903 | 20.7 |
| **Child sex** |  |  |
| Female | 2,142 | 49.1 |
| Male | 2,219 | 50.9 |
| **Birth order** |  |  |
| 1 | 1,521 | 34.9 |
| 2-3 | 1,840 | 42.2 |
| 4-5 | 699 | 16.0 |
| 6+ | 301 | 6.9 |

### Maternal Functional Difficulty

The weighted prevalence of severe maternal functional difficulty was 3.74% (SE 0.35). Table 2 shows the prevalence by domain. Hearing difficulty was the most common (2.20%), followed by difficulty remembering or concentrating (1.61%) and self-care (0.21%). No mothers in the analytic sample reported severe difficulty in seeing, walking, or communicating. The prevalence of any functional difficulty (moderate or severe) among all eligible women was 13.4%.

**Table 2.** Weighted prevalence of maternal functional difficulty and child undernutrition.

| Indicator | Weighted % | SE |
| --- | --- | --- |
| Maternal functional difficulty (severe, any domain) | 3.74 | 0.35 |
| **By domain:** |  |  |
| Seeing | 0.00 | 0.00 |
| Hearing | 2.20 | 0.28 |
| Walking | 0.00 | 0.00 |
| Remembering/concentrating | 1.61 | 0.23 |
| Self-care | 0.21 | 0.08 |
| Communicating | 0.00 | 0.00 |
| Child stunting (HAZ <-2 SD) | 36.4 | 1.15 |
| Mother: no difficulty | 36.4 | 1.16 |
| Mother: functional difficulty | 35.1 | 4.92 |
| Child underweight (WAZ <-2 SD) | 8.7 | 0.65 |
| Mother: no difficulty | 8.6 | 0.66 |
| Mother: functional difficulty | 10.4 | 2.93 |
| Child wasting (WHZ <-2 SD) | 1.7 | 0.27 |
| Mother: no difficulty | 1.7 | 0.27 |
| Mother: functional difficulty | 1.8 | 1.13 |

### Child Undernutrition

Overall, 36.4% of children were stunted (SE 1.15%), 8.7% were underweight (SE 0.65%), and 1.7% were wasted (SE 0.27%). The prevalence of stunting rose sharply with child age, from 16% in infants 0-11 months to over 45% in children 24-47 months. Male children had slightly higher wasting prevalence (2.2%) than female children (1.3%). Children of mothers with functional difficulty had similar stunting prevalence (35.1%) and slightly higher underweight prevalence (10.4%) compared to children of mothers without functional difficulty (36.4% and 8.6% respectively), though these differences were not statistically significant.

### Multivariable Logistic Regression

After adjustment for maternal age, education, height, wealth, residence, child age, child sex, and birth order, maternal functional difficulty was not associated with any of the three undernutrition outcomes (Table 3). The adjusted odds ratios were close to the null for stunting (AOR 0.85, 95% CI: 0.55-1.33), underweight (AOR 1.06, 95% CI: 0.55-2.02), and wasting (AOR 0.96, 95% CI: 0.26-3.53).

**Table 3.** Association between maternal functional difficulty and child undernutrition: adjusted odds ratios.

| Variable | Stunting AOR (95% CI) | Underweight AOR (95% CI) | Wasting AOR (95% CI) |
| --- | --- | --- | --- |
| **Maternal functional difficulty (ref: No)** |  |  |  |
| Yes | 0.85 (0.55-1.33) | 1.06 (0.55-2.02) | 0.96 (0.26-3.53) |
| **Maternal age (ref: 15-19)** |  |  |  |
| 20-29 | 1.07 (0.86-1.34) | 0.92 (0.62-1.37) | 1.09 (0.50-2.38) |
| 30-39 | 0.99 (0.78-1.24) | 1.11 (0.76-1.63) | 1.10 (0.53-2.30) |
| 40-49 | 1.04 (0.74-1.46) | 0.94 (0.50-1.78) | 2.20 (0.79-6.10) |
| **Maternal education (ref: Higher)** |  |  |  |
| No education | 1.34 (0.61-2.94) | 0.81 (0.22-2.90) | 0.61 (0.05-6.90) |
| Primary | 1.41 (0.70-2.88) | 0.64 (0.20-2.11) | 0.64 (0.07-5.82) |
| Secondary | 1.07 (0.52-2.22) | 0.43 (0.13-1.38) | 0.70 (0.08-6.15) |
| **Maternal height (ref: <145 cm)** |  |  |  |
| 145-149 cm | 0.69 (0.37-1.32) | 0.73 (0.36-1.49) | 0.73 (0.16-3.38) |
| ≥150 cm | 0.29 (0.16-0.54) | 0.36 (0.19-0.68) | 0.41 (0.10-1.72) |
| **Wealth (ref: Richest)** |  |  |  |
| Richer | 0.70 (0.49-0.99) | 0.72 (0.40-1.32) | 0.27 (0.09-0.87) |
| Middle | 1.05 (0.77-1.42) | 1.27 (0.74-2.18) | 0.65 (0.27-1.56) |
| Poorer | 1.14 (0.85-1.53) | 1.61 (0.97-2.70) | 0.81 (0.35-1.86) |
| Poorest | 1.05 (0.77-1.42) | 1.27 (0.74-2.18) | 0.65 (0.27-1.56) |
| **Residence (ref: Rural)** |  |  |  |
| Urban | 1.25 (0.92-1.70) | 1.31 (0.83-2.07) | 1.05 (0.44-2.53) |
| **Child age (ref: 0-11 mo)** |  |  |  |
| 12-23 | 2.16 (1.59-2.92) | 2.40 (1.48-3.90) | 1.33 (0.56-3.13) |
| 24-35 | 3.25 (2.38-4.43) | 1.62 (0.95-2.76) | 0.84 (0.32-2.21) |
| 36-47 | 3.04 (2.24-4.12) | 1.81 (1.03-3.19) | 0.56 (0.19-1.70) |
| 48-59 | 2.32 (1.69-3.19) | 2.07 (1.21-3.52) | 0.36 (0.12-1.10) |
| **Child sex (ref: Female)** |  |  |  |
| Male | 1.18 (0.99-1.41) | 1.07 (0.79-1.46) | 1.82 (0.95-3.49) |
| **Birth order (ref: 1)** |  |  |  |
| 2-3 | 0.95 (0.78-1.15) | 1.25 (0.90-1.74) | 0.88 (0.47-1.65) |
| 4-5 | 0.88 (0.67-1.15) | 1.18 (0.77-1.82) | 0.82 (0.41-1.66) |
| 6+ | 0.85 (0.58-1.26) | 1.21 (0.69-2.12) | 0.62 (0.21-1.83) |
| PAF (%) | -0.64% | 0.19% | -0.15% |

Note: AOR = adjusted odds ratio; CI = confidence interval; PAF = population attributable fraction. All models account for complex survey design. Marital status excluded (all mothers were married).

Several covariates showed strong independent associations with undernutrition. Child age was the strongest predictor of stunting: children aged 24-35 months had more than three times the odds of stunting compared to infants (AOR 3.25, 95% CI: 2.38-4.43). Maternal height of at least 150 cm was strongly protective against both stunting (AOR 0.29, 95% CI: 0.16-0.54) and underweight (AOR 0.36, 95% CI: 0.19-0.68). Children in the richest wealth quintile had lower odds of stunting (AOR 0.61, 95% CI: 0.42-0.89) and underweight (AOR 0.50, 95% CI: 0.26-0.98) compared to the poorest quintile.

### Population Attributable Fraction

The population attributable fractions for maternal functional difficulty were negligible: −0.64% for stunting, 0.19% for underweight, and −0.15% for wasting. These values suggest that even if a causal relationship were assumed, eliminating maternal functional difficulty would not meaningfully change the population prevalence of child undernutrition in Malawi.

### Sensitivity Analyses

Using the alternative “any difficulty” definition (moderate or severe), the results remained consistent with the primary analysis, with no significant associations observed for any of the three outcomes. Restricting the analysis to firstborn children or to children aged 12-59 months did not materially change the findings (results available upon request).

## Discussion

In a nationally representative sample of mother-child dyads in Malawi, we found no association between maternal functional difficulty and child stunting, underweight, or wasting. The adjusted odds ratios for the association were near the null for all three outcomes, and the population attributable fractions were negligible. This study is, to our knowledge, the first to investigate this association in sub-Saharan Africa.

Our findings differ from the results of research in Bangladesh using Multiple Indicator Cluster Survey 2019 data that reported the functional difficulty of the mother associates with moderate and severe malnutrition of boys ^10^. This discrepancy may be explained through different reasons. First, the prevalence of maternal functional difficulty in our study in Malawi (3.7%) is low compared to Bangladesh which could affect the power of our statistical association to pick up a small correlation. The second reason is related to cultural factors that are discussed later in the submission. Malawi’s community clearly upholds extended family caregiving.

Alternative caregivers such as grandmothers, aunts, and older siblings play an active role in child feeding and care^13^. This may explain compensating for the mother’s functional difficulty and allowing her child to have adequate nutritional status. In Bangladesh, nuclear families are common. Therefore, there is no support with feeding activities to the child when the mother could not do so due to her disability. The third reason is related to community support for nutrition programs in Malawi. Malawi practice community nutrition programs including growth monitoring and promotion sessions through surveillance health assistants and it proved implementing safety nets for children of disabled mothers^3^.

As for the characteristics of functional difficulties, it is interesting to note that hearing (2.2%) and cognitive (1.6%) disabilities are most prevalent in our study sample. These types of functional impairments are less likely to interfere with the actual physical care-related tasks of preparing children’s food and feeding them as are difficulties arising from mobility or self-care (neither of which were reported frequently by our sample). If it is the case that the most frequent functional difficulties experienced by caregivers of children with disabilities do not seriously limit these important child nutrition care activities, this may help explain the lack of significant findings in this study.

The positive association of maternal height with reduced odds of child stunting and underweight is consistent with ample evidence across various contexts^16,17^. As a measure of the intergenerational nutritional status and genetic factors which govern fetal growth and growth of child height ^17^, maternal height has important implication on child nutrition as well. The risk of stunting of children born to women who are shorter than 145cm is more than 3 times which reinforces the neglected importance of adolescent nutrition on birth outcome and further childhood undernutrition.

The expected increase in stunting with child age also shows the rise, from 16% in infants to more than 45% in 24-47 months aged children, is similar to globally observed patterns of growth faltering during the first two years of age ^18^. Child growth faltering during the early part of life is the effect of inadequately low intakes, recurrent illnesses and poor feeding practices during the most rapid growth period. Our results suggested the need to advocate nutrition interventions requiring higher intensity efforts during the first 1,000 days (conception to 2 years).

### Strengths

This study has several strengths. The facts derived from the data is nationally representative as it is obtained from a large survey designed with strict quality assurance techniques. The Washington Group Short Set is a validated, standardized tool for the assessment of disability, allowing international comparability to other studies. A complex sampling design was taken into consideration and all prespecified confounders that are derived from literature were accounted for.

### Limitations

Several limitations should be acknowledged. The cross-sectional design precludes causal inference; we cannot determine whether maternal functional difficulty preceded the development of child undernutrition. The low prevalence of maternal functional difficulty (3.7%), while consistent with other DHS-based studies from the region, may have resulted in insufficient statistical power to detect modest associations. This was particularly evident for the wasting outcome, where the confidence interval was wide (0.26–3.53). The Washington Group Short Set measures self-reported functional difficulty, which may be subject to social desirability or cultural reporting bias. We were unable to examine specific disability domains separately due to small cell sizes. All mothers in our analytic sample were married, reflecting the cultural context in Malawi, but this precluded examination of marital status as a potential effect modifier. We could not control for unmeasured confounders such as household food insecurity, child morbidity, or maternal mental health, which may influence both maternal disability reporting and child nutritional status. Finally, the “some difficulty” group was excluded from the primary analysis to maintain a clear contrast, which may have reduced generalizability.

### Policy implications

The policy implications arising from our results are noteworthy. The lack of association between maternal functional difficulty and child undernutrition should in no way imply that mothers with disabilities do not experience hardships. Rather, it implies that in the specific setting of Malawi, the existing systems of familial support and community integration may be shielding children from the risks posed by the maternal disabilities on nutritional outcomes. This is a positive finding. At the same time, it does not imply that mothers with disabilities do not require targeted interventions. These mothers may experience barriers to accessing health services, education, and livelihood opportunities^8^, and disability-inclusive health programming will remain impactful. Our findings only imply that maternal disability status should not be prioritized over other factors as an inclusion criterion for targeting interventions aimed at improving child nutrition in Malawi.

## Conclusion

Maternal functional difficulty was not associated with child stunting, underweight, or wasting in this nationally representative sample from Malawi. Population attributable fractions were negligible. These findings suggest that existing family and community support structures in Malawi may be effectively protecting children from the nutritional risks that could be expected to accompany maternal disability. Efforts to reduce child undernutrition should continue to prioritise maternal nutritional status (as reflected in maternal height), household economic conditions, and intensive interventions during the first 1,000 days of life.

## Data Availability Statement

The data that support the findings of this study are available from The DHS Program upon registration and approval of a concept paper. Restrictions apply to the availability of these data, which were used under license for this study. Data are available at https://www.dhsprogram.com.

## Declarations

The authors declare that this research has not been published in any other journal, therefore it is declared to comply with PLOS ONE publications policy. There are no images used, or other materials which require anonymity, the questionnaire followed informed consent and anonymity.

## Data usage Approval letter

The approval letter for DHS data use is available.

## Patient and Public Involvement

No patient or the public was involved in the design and conduct of this study

## Consent for publication

Not Applicable.

## Author’s contributions

Conceptualization-GCS; Methodology-RK & GCS,MMA,GM; Data preparation and formal analysis: GCS; Writing of Original draft – RK,GCS,NCN, BKK; Writing, review & editing –,NCN,SS, BKK,SK; Supervision & Validation; GCS,RK

## Acknowledgment

We would like to thank the DHS program for the data.

## Ethical approval

Not required.

## REFERENCES

1. Levels and trends in child malnutrition: UNICEF/WHO/World Bank Group joint child malnutrition estimates: key findings of the 2023 edition. https://www.who.int/publications/i/item/9789240073791.

2. Black, R. E. et al. Maternal and child undernutrition and overweight in low-income and middle-income countries. Lancet 382, 427–451 (2013).

3. National Statistical Office & Zomba, Malawi. Malawi Demographic and Health Survey. (2024).

4. Global nutrition targets 2025: stunting policy brief. https://www.who.int/publications/i/item/WHO-NMH-NHD-14.3.

5. World Report on Disability. https://www.who.int/teams/noncommunicable-diseases/sensory-functions-disability-and-rehabilitation/world-report-on-disability.

6. Madans, J. H., Loeb, M. E. & Altman, B. M. Measuring disability and monitoring the UN Convention on the Rights of Persons with Disabilities: the work of the Washington Group on Disability Statistics. BMC Public Health 11 Suppl 4, S4 (2011).

7. Groce, N., Bailey, N., Lang, R., Trani, J. F. & Kett, M. Water and sanitation issues for persons with disabilities in low- and middle-income countries: a literature review and discussion of implications for global health and international development. J Water Health 9, 617–627 (2011).

8. Mitra, S., Posarac, A. & Vick, B. Disability and Poverty in Developing Countries: A Multidimensional Study. World Development 41, 1–18 (2013).

9. Andrews, E. E., Powell, R. M. & Ayers, K. B. Experiences of Breastfeeding among Disabled Women. Womens Health Issues 31, 82–89 (2021).

10. Haq, I. et al. Gender differences in child nutrition status of Bangladesh: a multinomial modeling approach. Journal of Humanities and Applied Social Sciences 4, 379–392 (2021).

11. Anjum, A., Ahammed, T., Hasan, M. M., Chowdhury, M. A. B. & Uddin, M. J. Mother’s functional difficulty is affecting the child functioning: Findings from a nationally representative MICS 2019 cross-sectional survey in Bangladesh. Health Sci Rep 6, e1023 (2023).

12. The DHS Program - Methodology. https://dhsprogram.com/methodology/.

13. Heidkamp, R. A. et al. Mobilising evidence, data, and resources to achieve global maternal and child undernutrition targets and the Sustainable Development Goals: an agenda for action. Lancet 397, 1400–1418 (2021).

14. Lumley, T. Analysis of Complex Survey Samples. Journal of Statistical Software 9, 1–19 (2004).

15. Rockhill, B., Newman, B. & Weinberg, C. Use and misuse of population attributable fractions. Am J Public Health 88, 15–19 (1998).

16. Addo, O. Y. et al. Maternal height and child growth patterns. J Pediatr 163, 549–554 (2013).

17. Kozuki, N. et al. Short Maternal Stature Increases Risk of Small-for-Gestational-Age and Preterm Births in Low- and Middle-Income Countries: Individual Participant Data Meta-Analysis and Population Attributable Fraction. J Nutr 145, 2542–2550 (2015).

18. Prendergast, A. J. & Humphrey, J. H. The stunting syndrome in developing countries. Paediatr Int Child Health 34, 250–265 (2014).

